# Interactions between mild traumatic brain injury and genetics perturb neuronal and glial pathways and networks relevant to learning and memory in ABCD study

**DOI:** 10.1101/2025.03.11.25323465

**Authors:** Michael Cheng, Melody Mao, Wenjing Meng, Florin Vaida, Joanna Jacobus, Emily Troyer, Everett L. Delfel, Emily L. Dennis, Elisabeth A. Wilde, Tracy Abildskov, Nicola L. de Souza, Jeffrey E. Max, Xia Yang

**Author notes:** co-corresponding author* Xia Yang Department of Integrative Biology & Physiology, University of California, Los Angeles, Los Angeles, CA 90095, USA Jeffrey E Max Department of Psychiatry, University of California, San Diego, San Diego, CA 92093, USA.

## Abstract

Mild traumatic brain injury (mTBI) disproportionately affects children and adolescents and has been associated with poorer neurocognitive performance, but the variability in acute and chronic symptoms presents challenges in understanding the biological mechanisms underlying symptom heterogeneity and predicting these effects in clinical settings. We hypothesized that genetic factors interact with mTBI to determine vulnerability or resistance to neurological dysfunction post-mTBI. We leveraged the baseline Adolescent Brain Cognitive Development (ABCD) cohort to conduct a gene-by-mTBI genome-wide association study (GWAS) to study the interaction between mTBI and genetics in learning and memory compared to orthopedic injury controls. The GWAS revealed significant biological pathways involved in mitochondrial function and synaptic signaling that are enriched for SNPs showing evidence of interaction with mTBI. Integration of the gene-by-mTBI pathways from ABCD with cell-type specific gene regulatory networks built from single-cell RNA sequencing data from the Allen Brain Atlas uncovered key driver genes such as *APP*, *MAPT*, and *MOG* which coordinate between cell types in hippocampus and cortex to regulate these pathways. Lastly, we performed polygenic risk score (PRS) analysis on these pathways to assess their clinical value in predicting learning and memory outcomes in the ABCD cohort, revealing a statistically significant contribution but limited clinical benefit. Our findings provide novel insights into the genetic modifiers of mTBI pathology and propose potential therapeutic candidates at pathway and network levels.

**Author Summary:** Mild traumatic brain injury (mTBI), or concussion, is prevalent in adolescents and can have lasting impact on brain development, learning, and memory. However, the high variability in injury outcomes presents major challenges in predicting the specific recovery trajectories in individual children. Our study examines the entire genome to uncover genetic factors underlying mTBI response that determine an individual’s vulnerability to cognitive deficits. By investigating the interaction between genetics and injury, we aim to pinpoint how genetic predispositions affect biological processes in brain injury recovery to determine disease severity.

Our findings revealed certain genetic factors that are related to learning and memory in individuals with mTBI, but not in those with orthopedic injuries. These factors affect crucial areas of brain recovery, including neuronal repair and metabolism. We identified the core genes that coordinate across different brain cell types to affect these biological pathways. Finally, we leveraged these genetic factors to predict learning and memory performance in mTBI patients.

By examining the biological mechanisms driven by the genetic-mTBI interaction, we provide novel insights into the complex relationships between genetics, brain injury, and cognitive function. Our study provides a data-driven framework to understand how genetic and environmental factors interact to influence disease outcomes.

## Introduction

Traumatic brain injury is a global public health concern, with an estimated 69 million injuries occurring worldwide each year and 1.4 million reported annually in the United States alone [1,2]. Mild traumatic brain injury (mTBI), commonly known as concussion, is the most prevalent form of TBI, accounting for 70-90% of all cases [3]. Although “mild” suggests favorable prognosis, the effects of mTBI vary between individuals and can have lasting impacts on neural and psychological development [4–7]. With one-third of all mild TBIs occurring in children under the age of 15, accurate prognostication of injury outcomes in children and adolescents is necessary [8]. However, current mTBI prognostic models fail to capture the wide variation in patient outcomes [5,7], which is likely driven by the poorly understood genetic predisposition that interacts with mTBI to affect acute and chronic neurocognitive outcomes differently between individuals.

Recent efforts to explore genetic mechanisms of mTBI recovery have proposed candidate genetic variants affecting neuronal repair, inflammation, and mitochondrial dysfunction, such as *APOE*, *DRD2*, *COMT*, *BDNF*, and *BCL2*, through targeted genetic studies [9–19]. The *APOE4* risk allele has shown to negatively correlate with overall chronic TBI outcome as defined by the Glasgow Outcome Scale [20,21]. The *DRD2* variant is associated with poorer performance on the California Verbal Learning test for verbal learning and memory, and the *COMT* variant negatively correlates with the Wechsler Adult Intelligence Scale, fourth edition Processing Speed Index Subscale test for non-verbal processing speed [17,18]. With limited sample sizes between 40 and 100 patients, these studies resort to targeted genetic studies because of the lack of statistical power to conduct an unbiased genome-wide association study (GWAS). Thus, assessing the genome-wide interactions between genetic variants and mTBI and their clinical manifestations remains an ongoing effort. In addition to individual gene candidates, novel systems biology studies on pediatric patients with mTBI and orthopedic injury (OI) revealed marginal and aggregate associations of neuronal and inflammatory pathways with child behavioral adjustment and executive function specifically in the mTBI patients [17,22]. These studies explore a few predefined gene sets containing genes related to TBI in cohorts with sample sizes less than 100 patients, which may limit the possibility of discovering novel genetic pathways or genes. Moreover, there is a lack of data-driven genetic studies on learning and memory consequences to mTBI despite the well-known association between mTBI and cognitive performance to elucidate novel mechanisms.

The Adolescent Brain Cognitive Development (ABCD) cohort provides an opportunity to conduct GWAS on how mTBI interacts with genetic risks to affect neurological outcome [23,24]. With over 11,000 children enrolled in a ten-year longitudinal study that measures diverse clinical variables, brain scans, and genetic data, this study, while modestly sized, contains the largest available mTBI (n=423) and OI (n=1,469) populations to perform retrospective GWAS analyses. Understanding genetic mechanisms can be further bolstered by the advent of single cell RNA sequencing (scRNA-seq) from the human and mouse brain, such as the Allen Brain Atlas, to enable modeling of gene regulatory networks in different brain regions and cell types [25,26]. Integrating the genetic and transcriptomic layers provides more granularity to resolve the genetic contributions of specific molecular pathways and networks in individual brain cell types on mTBI clinical outcome. Our current study aims to leverage the abundant data from ABCD and Allen Brain Atlas to identify gene by mTBI interactions to influence learning and memory through brain cell type specific pathways and networks, and further test the clinical relevance of our findings using polygenic risk score analysis (Figure 1A).

**Fig 1:**
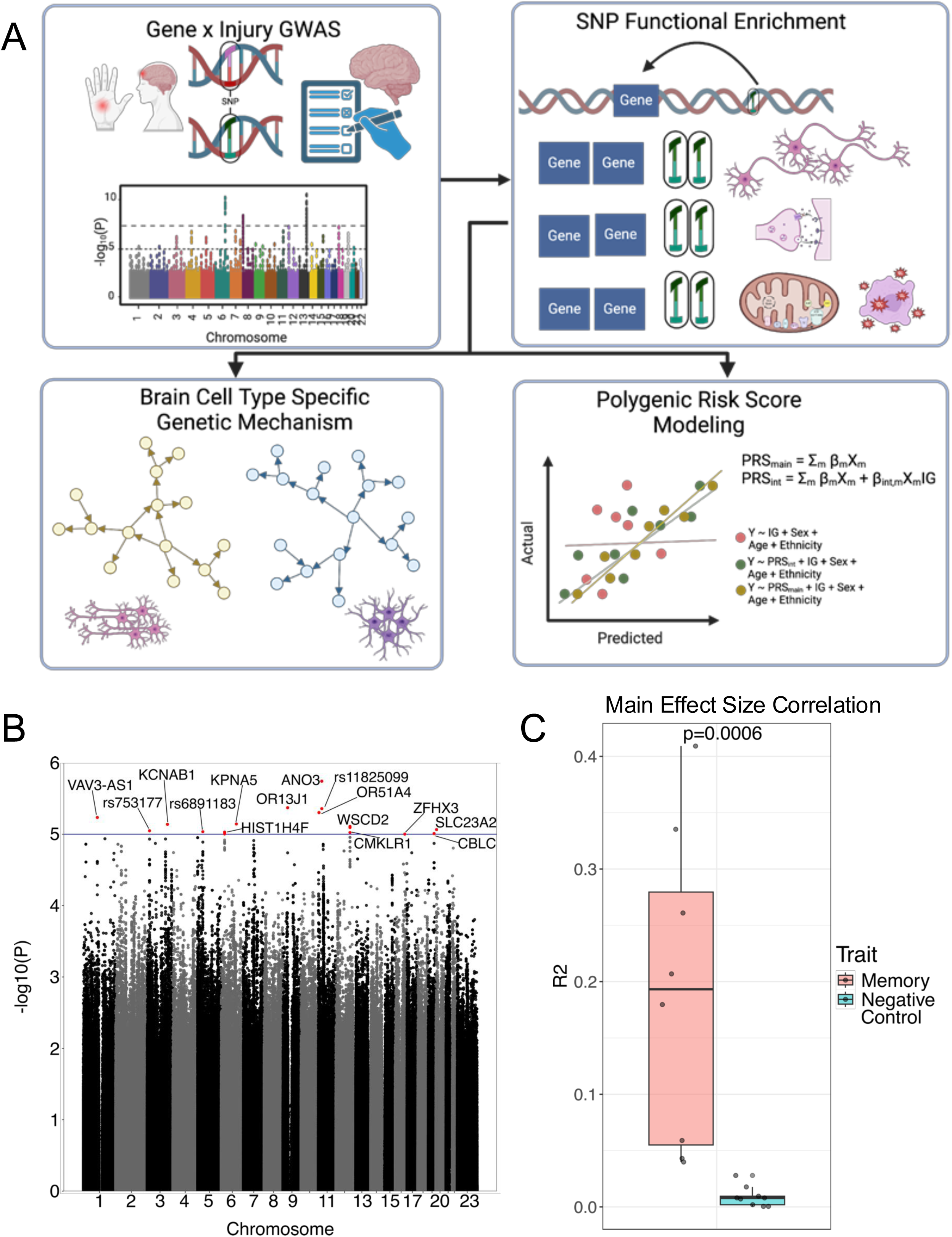
Overview and Validation with Additive model. A: Overview of GWAS and functional analysis. First a gene by environment (mTBI) GWAS on learning and memory composite scores was performed, followed by SNP to gene mapping and biological pathway enrichment of GWAS signals. These pathways are overlaid onto cell type gene regulatory networks for mechanistic study and curated into a polygenic risk score for clinical modeling. B: Manhattan Plot of GWAS additive model. Blue line is the genome wide suggestive threshold p=1e-5. C: Pearson strength of correlation boxplots between ABCD main-effect GWAS and previous GWAS. Red boxplot consists of working, verbal, and spatial memory GWAS effect sizes, which is expected to have higher effect size correlation. Blue boxplot consists of phenotypes unrelated to memory or learning, where SNP effect sizes are expected to be less correlated.

## Results

### Retrospective baseline data from ABCD

Of the 11,878 children enrolled in the ABCD project at baseline, 10,870 had available genotype data. Of the genotyped participants, 8,978 had no injury at baseline, 1,469 reported an OI, and 423 were classified as having an mTBI according to the Ohio State Traumatic Brain Injury Assessment [27,28] (details in Methods). After filtering the cohort to unrelated individuals based on genetic relatedness, 1,180 OI and 342 mTBI cases were included in the GWAS analysis.

To obtain learning and memory scores in the participants, we followed the Bayesian probabilistic principal component analysis conducted in Thompson et. al [29]. This method aggregates 9 administered test scores (NIH Toolbox measures Picture Vocabulary Task, Oral Reading Recognition Task, Pattern Comparison Processing Speed Test, List Sorting Working Memory Test, Picture Sequence Memory Test, Flanker Task, and Dimensional Change Card Sort Task; Rey Auditory Verbal Learning Task; and Little Man Task) into 3 neurocognitive principal components (NPCs) that correlate with general cognition (NPC1), executive function (NPC2), and learning and memory (NPC3). The learning and memory NPC3 contained the highest principal component loadings for the Toolbox Picture Sequence Memory Task, Toolbox List Sorting Working Memory Task, and Rey Auditory Verbal Learning Task, and was the primary focus of the current study. We chose to use the principal components instead of individual phenotypic traits to capture the overall learning and memory performance and to reduce multiple testing burden in this modestly sized cohort.

### Learning and memory NPC3 GWAS on mTBI and OI patients at baseline recapitulates main genetic effects from existing GWAS studies of working memory

To first assess if the ABCD cohort can capture genetic factors associated with learning and memory uncovered in previous GWAS studies, we carried out a main-effect GWAS focusing on the effect of SNPs on NPC3 without an interaction term. The covariates in the main effect GWAS included injury type (mTBI vs OI), age, sex, and top 20 genetic principal components representing the overall genetic architecture for population ancestry differences.

From the main-effect GWAS, there were 21 SNPs that passed the genome-wide suggestive p-value threshold of 1e-5, but no SNPs passed genome-wide significance below 5e-8 (Figure 1B; Supplementary Table 1). This is expected of the small sample size in the ABCD cohort. Nevertheless, the SNP effect size estimates from our ABCD GWAS correlate highly with previous working memory GWAS [30–34] with larger population sizes (450-50,000 individuals), and the correlations with previous working memory GWAS were significantly greater than those with unrelated phenotypes (fasting blood glucose [35], grip strength [36], low density lipoprotein cholesterol [37], ascending aortic diameter [38], hemoglobin levels [39], alkaline phosphatase [40], serum albumin levels [41], bone mineral density [42], triglyceride levels [43]) from the GWAS catalog (Figure 1C, p<0.001 by Wilcoxon rank sum test; Supplementary Table 2) [30–34]. These results support that the ABCD cohort recapitulates the main genetic effect of a similar cognitive memory trait, despite its smaller sample size, and provides confidence for conducting genetic-by-mTBI interaction analyses.

### SNP-by-mTBI interaction GWAS on learning and memory NPC3 reveal SNPs relevant to cognitive traits

To understand the presence of gene-injury interactions in association with learning, we added an interaction term between the SNP and injury type to account for interactive effects between mTBI (vs OI) and genetic variants into the above main effect GWAS, adjusting for the same covariates as above. The SNPs from the interaction term implicates genetic variants with differential effects on cognition between mTBI and OI.

In this interaction GWAS, 26 SNPs passed the suggestive threshold for the interaction term but no genome-wide significant signals were identified (Figure 2A; Supplementary Table 3). This again is due to the small sample size in ABCD, even though this is the largest cohort available to examine gene-by-mTBI interactions. To assess the validity and relevance of the GWAS signals from our ABCD study, we hypothesize that the genetic signals modifying mTBI effect should be relevant to cognitive traits as well. Therefore, we established two sets of positive control traits (50 in each set; should be relevant to mTBI and neurocognition related traits such as cognitive ability and household income) and two sets of negative control sets (50 in each set; should be less relevant to mTBI and neurocognition, such as aortic diameter or liver enzyme levels) by two independent neuroscientists (Supplementary Table 4). These traits were selected from all 2,891 traits collected in GWAS catalog [44]. SNPs associated with each of the positive and negative control traits were also extracted from the GWAS catalog. To test whether our SNP association signals from the SNP-by-mTBI interaction term are more enriched for the SNPs from the positive control GWAS traits than those from the negative control traits, we performed Mergeomics Marker Set Enrichment Analysis (MSEA) [45]. MSEA considers the whole spectrum of SNPs from our GWAS to include subtle and moderate genetic associations beyond the top genetic associations, and tests for enrichment of stronger association p-values among a given SNP set (here the SNPs from a positive control or a negative control trait) compared to randomly selected SNPs using a chi-like statistic summarized across various GWAS p-value cutoffs. The p-value distribution of the positive control traits is significantly greater than the negative control traits defined by both neuroscientists (Figure 2B, Wilcoxon rank sum test p=0.03 and p=0.002, respectively).

**Fig 2:**
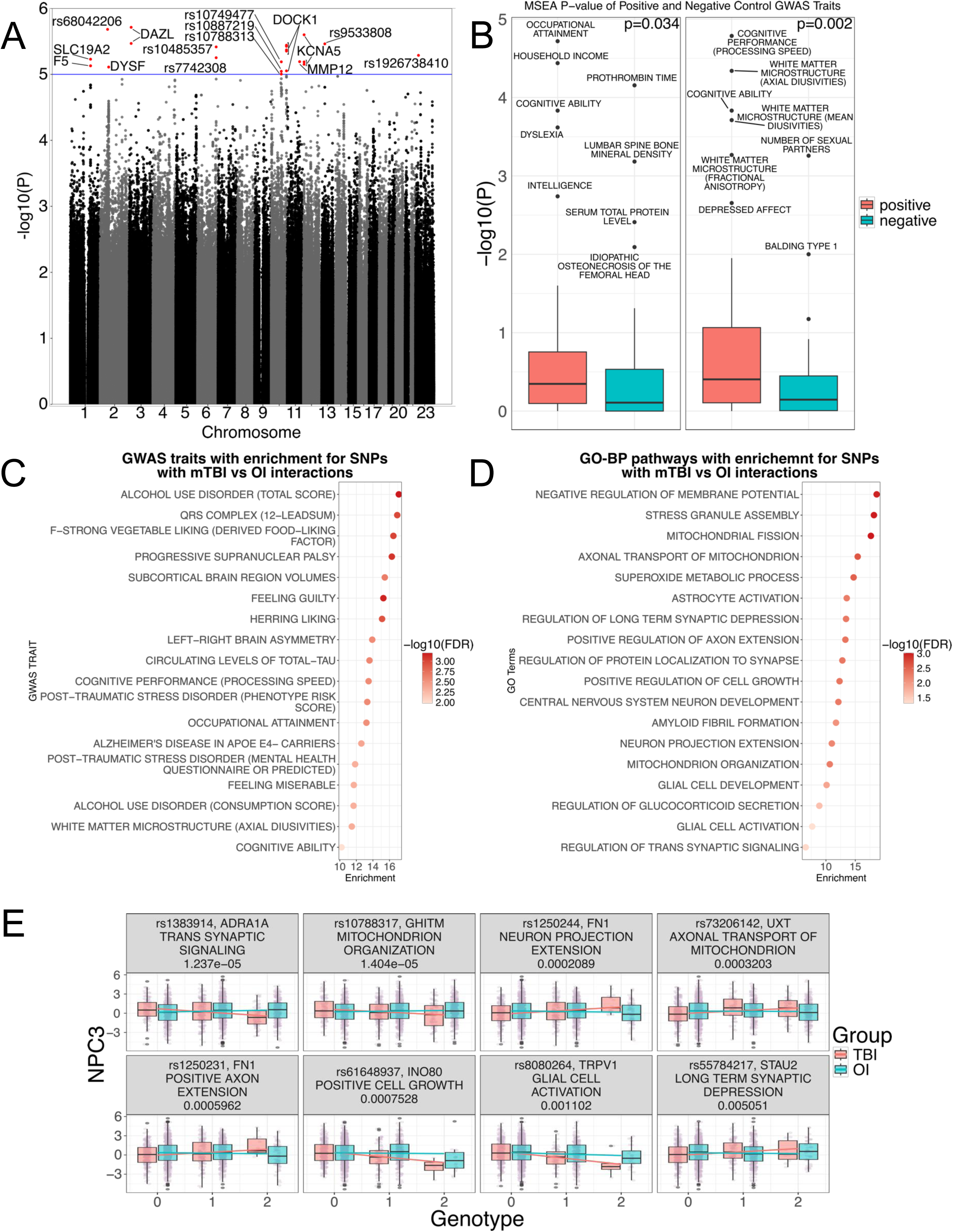
SNP-by-mTBI Interaction GWAS. A: Manhattan plot of the SNPxInjury term. Blue line is the genome wide suggestive threshold p=1e-5. B: Distribution of MSEA -log10(p-values) in positive and negative control GWAS traits from GWAS catalog determined by 2 neuroscientists. Traits from each neuroscientist is displayed separately. C: Significant marker set enrichment of top traits in the GWAS catalog. Color indicates FDR after BH correction. D: Significant marker set enrichment of top pathways in the Gene Ontology Biological Pathways. Color indicates FDR after BH correction. E: Learning and memory score distribution for top SNPs in GO pathways in Fig 2D. Color corresponds to injury group. Regression lines fit through linear regression.

We also tested all 2,891 GWAS traits in the GWAS catalog using MSEA, and 60 traits were significantly enriched for our NPC3 interaction SNPs after multiple testing correction. Among the 60 GWAS traits, 34 are related to mTBI, cognition and intelligence, such as cognitive ability, post-traumatic stress disorder, occupational attainment, household income, and dyslexia (Figure 2B, Supplementary Table 5). These results support that the genetic association signals from our SNP-by-mTBI interaction term were more similar to known mTBI and cognition related traits, thereby carrying biologically relevant information.

### SNP-by-mTBI Interaction GWAS on Learning and Memory NPC3 reveals significant biological pathways informed by SNPs with stronger associations

To interpret the functional implications of SNP-by-mTBI interaction SNPs using biological pathways, we performed the same MSEA analysis on 5,281 Gene Ontology biological pathways (GO-BP). We observed significant enrichment of our GWAS SNPs in 137 pathways out of 5,281 GO-BP pathways at false discovery rate (FDR) <5%, many of which were related to known TBI pathological mechanisms (Table 1, Supplementary Table 6). Pathways with the highest enrichments fell into broad categories of mitochondrial organization and metabolism, cytoskeletal organization and protein transport, neuronal development, synaptic signaling, and glial cell activation (Figure 2C).

**Table 1:** MSEA analysis of GO-BP pathways for enrichment of SNP-by-mTBI GWAS signals

| Pathway | FDR | MSEA Enrichment Score | Top Genes | Top SNPs in SNP-by-mTBI interaction term | -log10 (GWAS P) |
| --- | --- | --- | --- | --- | --- |
| MITOCHONDRIAL FISSION | 9.72E-04 | 17.65 | MAPT; AURKA; DDHD2; PINK1; FIS1 | rs17763658; rs59173918; rs72644707; rs12401770; rs150814238 | 3.77; 3.65; 3.54; 3.00; 2.80 |
| STRESS GRANULE ASSEMBLY | 1.01E-03 | 18.17 | MAPT; PRKAA2; BICD1; LSM14A; PUM2 | rs17763658; rs4568857; rs144166034; rs570643; rs74333499 | 3.77; 2.65; 2.53; 2.14; 2.11 |
| NEGATIVE REGULATION OF MEMBRANE POTENTIAL | 1.08E-03 | 18.63 | MAPT; TRPV1; PMAIP1; ARL6IP5; BAX | rs17763658; rs9905037; rs58697140; rs4407408; rs12460533 | 3.77; 3.62; 2.29; 2.26; 2.16 |
| AXONAL TRANSPORT OF MITOCHONDRION | 2.46E-03 | 15.40 | MAPT; TRAK1; OPA1; AGBL4; AGTPBP1 | rs17763658; rs9861303; rs6771174; rs320028; rs79634858 | 3.77; 2.75; 2.60; 2.25; 2.17 |
| SUPEROXIDE METABOLIC PROCESS | 3.03E-03 | 14.71 | MAPT; IMMP2L; NOX1; CYBA; ACP5 | rs17763658; rs17158597; rs7128; rs13334987; rs4804617 | 3.77; 3.25; 3.08; 3.06; 2.85 |
| POSITIVE REGULATION OF AXON EXTENSION | 4.51E-03 | 13.31 | TRPC5; MAPT; FN1; NTN1; DSCAM | rs3027784; rs17763658; rs1250244; rs62068228; rs7277154 | 3.77; 3.77; 3.68; 3.26; 2.78 |
| POSITIVE REGULATION OF CELL GROWTH | 4.61E-03 | 12.32 | SLC44A4; IST1; TRPC5; MAPT; FN1 | rs146535109; rs79169855; rs3027784; rs17763658; rs1250244 | 4.69; 3.88; 3.77; 3.77; 3.68 |
| REGULATION OF PROTEIN LOCALIZATION TO SYNAPSE | 4.74E-03 | 12.80 | MAPT; ARHGAP44; DVL1; GHSR; WNT7A | rs17763658; rs12950148; rs2296475; rs595161; rs11128663 | 3.77; 3.39; 2.87; 2.83; 2.72 |
| REGULATION OF LONG TERM SYNAPTIC DEPRESSION | 4.98E-03 | 13.42 | MAPT; GRID2; ADCY8; STAU2; FMR1 | rs17763658; rs17274136; rs191736; rs55784217; rs25726 | 3.77; 2.57; 2.54; 2.30; 2.20 |
| CENTRAL NERVOUS SYSTEM NEURON DEVELOPMENT | 5.20E-03 | 12.12 | ZMIZ1; MAPT; NDNF; DRD1; NIN | rs704016; rs17763658; rs17051286; rs267373; rs2984270 | 4.87; 3.77; 3.47; 3.41; 3.41 |
| ASTROCYTE ACTIVATION | 5.41E-03 | 13.54 | MAPT; IL1B; IFNGR1; LDLR; MT1X | rs17763658; rs1143629; rs4896242; rs379309; rs12934069 | 3.77; 2.52; 2.37; 2.26; 2.20 |
| MITOCHONDRION ORGANIZATION | 5.44E-03 | 10.65 | GHITM; HSPA1A; HSPA1L; ERBB4; MAN2A1 | rs10788317; rs146535109; rs146535109; rs2165098; rs2033463 | 4.85; 4.69; 4.69; 4.15; 3.77 |
| NEURON PROJECTION EXTENSION | 6.75E-03 | 11.00 | TRPC5; MAPT; FN1; AURKA; CPNE5 | rs3027784; rs17763658; rs1250244; rs59173918; rs6920224 | 3.77; 3.77; 3.68; 3.65; 3.62 |
| AMYLOID FIBRIL FORMATION | 8.83E-03 | 11.71 | MAPT; GSN; SIAH2; IAPP; RIPK3 | rs17763658; rs2304392; rs73869296; rs28446944; rs17256902 | 3.77; 3.12; 2.92; 2.83; 2.65 |
| GLIAL CELL DEVELOPMENT | 1.31E-02 | 10.11 | MAPT; ASPA; SOX11; DRD1; CDK6 | rs17763658; rs9905037; rs76251724; rs267373; rs11974095 | 3.77; 3.62; 3.42; 3.41; 3.23 |
| REGULATION OF GLUCOCORTICOID SECRETION | 2.42E-02 | 8.87 | CRHR1; TSPO; GAL; POMC; GALR1 | rs7222658; rs138952; rs59226850; rs3754863; rs2849291 | 3.99; 2.67; 2.27; 1.77; 1.68 |
| REGULATION OF TRANS SYNAPTIC SIGNALING | 4.18E-02 | 6.55 | DYSF; ADRA1A; PLCB1; ERC2; SYN1 | rs4852258; rs1383914; rs75234999; rs56242225; rs4824626 | 5.11; 4.91; 4.41; 4.36; 3.99 |
| GLIAL CELL ACTIVATION | 4.92E-02 | 7.66 | MAPT; TRPV1; IL33; TYROBP; SNCA | rs17763658; rs9905037; rs16924291; rs4806237; rs200955273 | 3.77; 3.62; 3.14; 2.74; 2.52 |

To visualize the interaction effect, we plotted the SNPs with the strongest p-values from the interaction GWAS within the top significant pathways. The genotypes of these SNPs indeed show differential association with learning memory NPC3 scores between the ABCD mTBI and OI groups (Figure 2D), where SNPs show association with NPC3 in mTBI but not in OI.

The most significant SNPs whose minor alleles show negative associations with learning and memory specifically in mTBI patients, rs1383914 and rs10788317, map to the synaptic signaling gene alpha 1-adrenergic receptor (*ADRA1A*) and the growth hormone inducible transmembrane protein (*GHITM*, also known as TMBIM5 and MICS1 at protein level) in the mitochondrial organization pathway, respectively. *ADRA1A* has been linked to working memory dysfunction in TBI rat models [46]. The SNP specifically has been linked to the incidence and impact of fibromyalgia, a chronic pain disorder related to common pain-related symptoms post-mTBI and burdens on working memory capacity [47–51]. *GHITM* is responsible for mitochondrial formation and calcium ion regulation to maintain cell metabolism and survival. It does this through binding to and regulating the activity of cytochrome c along with Parkinson’s disease-associated intermembrane mitochondrial protein CHCHD2 [52,53]. On the other hand, the minor alleles of rs1250244 and rs1250231, which map to the fibronectin gene Fibronectin 1 (*FN1*), demonstrate positive association with learning specifically in the mTBI group but no association in the OI group. FN1 is a major downstream effector of transcription factor 4 in the cortex and hippocampus to influence neuronal positioning in development and spatial learning and reasoning [54]. It has been shown to play an important role in mTBI recovery as well, thereby carrying promise as a potential TBI therapeutic target [55,56]. Plasma fibronectin demonstrated neuroprotective effects following TBI, with mice deficient in the protein performing significantly worse in motor and cognitive tasks [57]. These SNPs did not show association with NPC3 in the OI group. Altogether, our findings support that the top SNPs relating to synaptic signaling, neuronal projection, and mitochondrial function interact with mTBI, but not OI, to affect learning and memory.

### Cell-type-specific network analysis reveals key regulators and networks governing neuronal repair pathways in the hippocampus and prefrontal cortex

To dissect the cell types and gene regulatory networks (GRNs) responsible for the significant pathways enriched for SNP-by-injury interaction in relevant brain regions, we constructed cell-type specific GRNs and performed Mergeomics Key Driver Analysis (KDA) [45] on the significant mTBI-mediated genetic pathways. We inferred cell-type-specific GRNs from scRNA-seq data from the mouse hippocampus and cortex in the Allen Brain Atlas [26] to elucidate the gene regulatory programs that define each cell type using SCING, a gradient-boosting and bootstrapping based global GRN construction method [58]. KDA employs a permutation approach on the GRN topology to identify hub genes, or key drivers, that are highly connected to genes in the biological pathways of interest. We performed KDA on the enriched GO-BP pathways using cell type GRNs from the hippocampus and various cortical regions (prelimbic, infralimbic, and orbital cortex; frontal pole and secondary motor cortex; and anterior cingulate cortex) (Table 2, Supplementary Table 7). We focused on these brain regions due to their relevance to TBI.

**Table 2:** Top key drivers for the significant mTBI-interaction pathways in hippocampal and cortical cell types

| Key Driver | Cell Type | Pathway | KDA FDR | KDA Enrichment | Top SNPs in SNP-by-mTBI interaction term | -log<sub>10</sub> (GWAS P) |
| --- | --- | --- | --- | --- | --- | --- |
| **Hippocampus** |  |  |  |  |  |  |
| CPLX2 | excitatory | REGULATION OF TRANS SYNAPTIC SIGNALING | 9.06E-06 | 9.6 | rs13174727; rs11134935; rs999065; rs11134936; rs13158114 | 1.06e-03; 1.27e-03; 1.45e-03; 6.12e-03; 6.88e-03 |
| PJA2 | excitatory | REGULATION OF TRANS SYNAPTIC SIGNALING | 6.31E-05 | 8.03 | rs9326769; rs1594076; rs2416213; rs7705657; rs2914711 | 2.38e-03; 4.42e-03; 4.45e-03; 7.03e-03; 7.47e-03 |
| MAPT | excitatory | REGULATION OF TRANS SYNAPTIC SIGNALING | 1.00E-02 | 5.63 | rs17763658; rs9890016; rs73313152; rs9896752; rs62061733 | 1.71e-04; 1.57e-03; 4.05e-03; 8.59e-03; 2.69e-02 |
| APP | excitatory | REGULATION OF TRANS SYNAPTIC SIGNALING | 3.13E-02 | 5.63 | rs440666; rs71317439; rs71317438; rs1580939; rs2211769 | 1.51e-02; 2.47e-02; 2.95e-02; 2.96e-02; 2.99e-02 |
| YWHAG | excitatory | REGULATION OF TRANS SYNAPTIC SIGNALING | 3.13E-02 | 6.5 | rs2868371; rs2908195; rs2908203; rs2070804; rs62476798 | 1.32e-02; 5.99e-02; 8.38e-02; 9.58e-02; 1.37e-01 |
| NDUFS6 | inhibitory | MITOCHONDRION ORGANIZATION | 9.58E-08 | 9.72 | rs6859291; rs904756; rs6555014; rs4296839; rs16884543 | 1.15e-02; 6.50e-02; 8.12e-02; 8.23e-02; 8.53e-02 |
| COX5A | inhibitory | MITOCHONDRION ORGANIZATION | 3.12E-04 | 11.41 | rs11638576; rs78324724; rs8033883; rs7175135; rs6495129 | 1.27e-01; 1.30e-01; 1.79e-01; 1.79e-01; 2.36e-01 |
| NDUFC1 | inhibitory | MITOCHONDRION ORGANIZATION | 4.45E-04 | 7.02 | rs34711204; rs6814602; rs6536006; rs1901173; rs7682630 | 6.72e-03; 4.00e-02; 5.01e-02; 7.48e-02; 7.74e-02 |
| COX17 | inhibitory | MITOCHONDRION ORGANIZATION | 4.87E-04 | 8.3 | rs149967762; rs71325395; rs13076819; rs4687877; rs2280580 | 1.10e-02; 1.62e-02; 1.72e-02; 1.96e-02; 2.91e-02 |
| NDUFA7 | inhibitory | MITOCHONDRION ORGANIZATION | 6.51E-04 | 8.33 | rs448652; rs7248963; rs12982876; rs34661506; rs112110995 | 8.31e-02; 1.01e-01; 1.02e-01; 1.02e-01; 1.02e-01 |
| PSAP | inhibitory | REGULATION OF TRANS SYNAPTIC SIGNALING | 5.76E-03 | 12.13 | rs9415055; rs9415050; rs118122154; rs7092990; rs115467618 | 3.43e-03; 4.52e-03; 1.69e-02; 2.23e-02; 2.44e-02 |
| TSPAN2 | oligodendrocyte | GLIAL CELL DEVELOPMENT | 6.56E-03 | 36.74 | rs6537851; rs74113933; rs12132759; rs6679900; rs3820630 | 3.95e-02; 8.14e-02; 9.05e-02; 9.99e-02; 1.05e-01 |
| MOG | oligodendrocyte | GLIAL CELL DEVELOPMENT | 1.57E-02 | 22.61 | rs9258119; rs9257928; rs29273; rs29270; rs45617437 | 3.98e-02; 5.18e-02; 6.15e-02; 6.99e-02; 6.99e-02 |
| CLDN11 | oligodendrocyte | GLIAL CELL DEVELOPMENT | 2.12E-02 | 36.74 | rs56226070; rs2901623; rs4955726; rs13434085; rs34814917 | 2.83e-03; 1.43e-02; 2.53e-02; 2.84e-02; 3.49e-02 |
| CNP | oligodendrocyte | GLIAL CELL DEVELOPMENT | 2.12E-02 | 40.08 | rs11079027; rs57825218; rs199922737; rs62076888; rs78071022 | 5.74e-02; 6.59e-02; 7.07e-02; 8.91e-02; 1.06e-01 |
| **Anterior Cingulate Cortex** |  |  |  |  |  |  |
| GRM7 | excitatory | REGULATION OF TRANS SYNAPTIC SIGNALING | 6.72E-03 | 10.38 | rs712793; rs712774; rs34735406; rs712783; rs41361551 | 9.94e-04; 1.39e-03; 3.65e-03; 4.92e-03; 6.38e-03 |
| NSF | excitatory | REGULATION OF TRANS SYNAPTIC SIGNALING | 6.72E-03 | 10.92 | rs199496; rs929254; rs199528; rs74939963; rs113839801 | 5.33e-03; 1.39e-02; 4.13e-02; 4.36e-02; 4.45e-02 |
| GRM5 | excitatory | REGULATION OF TRANS SYNAPTIC SIGNALING | 1.22E-02 | 15.38 | rs308804; rs1552739; rs56859701; rs973716; rs11826393 | 1.07e-04; 4.06e-04; 4.08e-04; 4.08e-04; 4.14e-04 |
| SV2A | inhibitory | REGULATION OF TRANS SYNAPTIC SIGNALING | 3.23E-05 | 7.52 | rs6587723; rs6696191; rs577935; rs626785; rs147211319 | 5.88e-02; 9.06e-02; 1.02e-01; 1.43e-01; 1.61e-01 |
| GRM5 | inhibitory | REGULATION OF TRANS SYNAPTIC SIGNALING | 6.53E-05 | 13.27 | rs308804; rs1552739; rs56859701; rs973716; rs11826393 | 1.07e-04; 4.06e-04; 4.08e-04; 4.08e-04; 4.14e-04 |
| NSF | inhibitory | REGULATION OF TRANS SYNAPTIC SIGNALING | 3.62E-02 | 9.34 | rs199496; rs929254; rs199528; rs74939963; rs113839801 | 5.33e-03; 1.39e-02; 4.13e-02; 4.36e-02; 4.45e-02 |
| CHL1 | oligodendrocyte | REGULATION OF TRANS SYNAPTIC SIGNALING | 9.76E-03 | 15.89 | rs4685713; rs11715727; rs9310805; rs13086886; rs77030744 | 4.75e-03; 1.39e-02; 2.11e-02; 2.29e-02; 2.54e-02 |
| TSPAN2 | oligodendrocyte | GLIAL CELL DEVELOPMENT | 9.76E-03 | 43.97 | rs6537851; rs74113933; rs12132759; rs6679900; rs3820630 | 3.95e-02; 8.14e-02; 9.05e-02; 9.99e-02; 1.05e-01 |
| **Prelimbic Infralimbic Orbital Cortex** |  |  |  |  |  |  |
| CX3CL1 | excitatory | REGULATION OF TRANS SYNAPTIC SIGNALING | 2.58E-03 | 11.49 | rs72784878; rs223880; rs148546737; rs4784801; rs757307 | 4.99e-02; 5.35e-02; 6.13e-02; 6.40e-02; 6.68e-02 |
| GRIN2A | excitatory | REGULATION OF TRANS SYNAPTIC SIGNALING | 8.37E-03 | 13.44 | rs7189919; rs8060390; rs56064109; rs7201288; rs8055473 | 1.14e-02; 1.93e-02; 1.94e-02; 2.21e-02; 2.25e-02 |
| MEF2C | inhibitory | REGULATION OF TRANS SYNAPTIC SIGNALING | 2.52E-02 | 11.65 | rs302482; rs302484; rs80043958; rs244752; rs244751 | 2.94e-02; 4.09e-02; 5.40e-02; 6.14e-02; 6.14e-02 |
| GRIA4 | inhibitory | REGULATION OF TRANS SYNAPTIC SIGNALING | 5.29E-04 | 9.65 | rs72979301; rs78442630; rs675091; rs17391463; rs10895891 | 1.24e-02; 2.15e-02; 3.37e-02; 5.37e-02; 5.59e-02 |
| ATP5PD | inhibitory | MITOCHONDRION ORGANIZATION | 5.94E-03 | 7.2 | None | None |
| MOG | oligodendrocyte | GLIAL CELL DEVELOPMENT | 4.82E-02 | 32.78 | rs9258119; rs9257928; rs29273; rs29270; rs45617437 | 3.98e-02; 5.18e-02; 6.15e-02; 6.99e-02; 6.99e-02 |
| **Frontal Pole Secondary Motor Cortex** |  |  |  |  |  |  |
| ITPR1 | excitatory | REGULATION OF TRANS SYNAPTIC SIGNALING | 9.57E-03 | 7.42 | rs6765702; rs73004812; rs2633717; rs73004818; rs73003098 | 1.91e-03; 2.86e-03; 3.19e-03; 3.37e-03; 3.51e-03 |
| NSF | excitatory | REGULATION OF TRANS SYNAPTIC SIGNALING | 9.93E-03 | 10.3 | rs199496; rs929254; rs199528; rs74939963; rs113839801 | 5.33e-03; 1.39e-02; 4.13e-02; 4.36e-02; 4.45e-02 |
| MOG | oligodendrocyte | GLIAL CELL DEVELOPMENT | 1.64E-02 | 31.18 | rs9258119; rs9257928; rs29273; rs29270; rs45617437 | 3.98e-02; 5.18e-02; 6.15e-02; 6.99e-02; 6.99e-02 |

Using the hippocampus cell type GRNs, we identified numerous key drivers at FDR<5% that regulated distinct processes in excitatory and inhibitory neurons, as well as in oligodendrocytes (Figure 3). In the excitatory neuron GRN, key drivers are involved in trans-synaptic signaling (e.g., *CPLX2;* FDR=9.06e-6). Numerous hub genes in this network are known to be associated with AD pathology (e.g. *MAPT, APP, PJA2, YWHAG*) (Figure 3A). *MAPT* (FDR=0.01) encodes the tau protein responsible for neurofibrillary tangles and *APP* (FDR=0.031) encodes the amyloid precursor protein that forms amyloid plaques in AD patients [59–61]. *PJA2* (FDR=6.31e-5), is essential for long term memory and synaptic plasticity, and inhibits *APP, MAPT*, and gamma-secretase activating protein *GSAP* expression in mice [62]. Also, *YWHAG* (FDR=0.031) is a biomarker for AD [63]. The inhibitory neuron key drivers mainly regulate the mitochondrial organization pathway with many KDs from the NADH dehydrogenase Complex 1 gene family, such as *NDUFS6* (FDR=9.58e-8), and the cytochrome c oxidation family like *COX5* (FDR=3.12e-4) (Figure 3C). Lastly, the oligodendrocyte network contains KDs promoting glial cell development, specifically through myelination genes *TSPAN2* (FDR=6.56e-3), *MOG* (FDR=0.016), *CLDN11* (FDR=0.021), and *CNP* (FDR=0.021) (Figure 3B).

**Fig 3:**
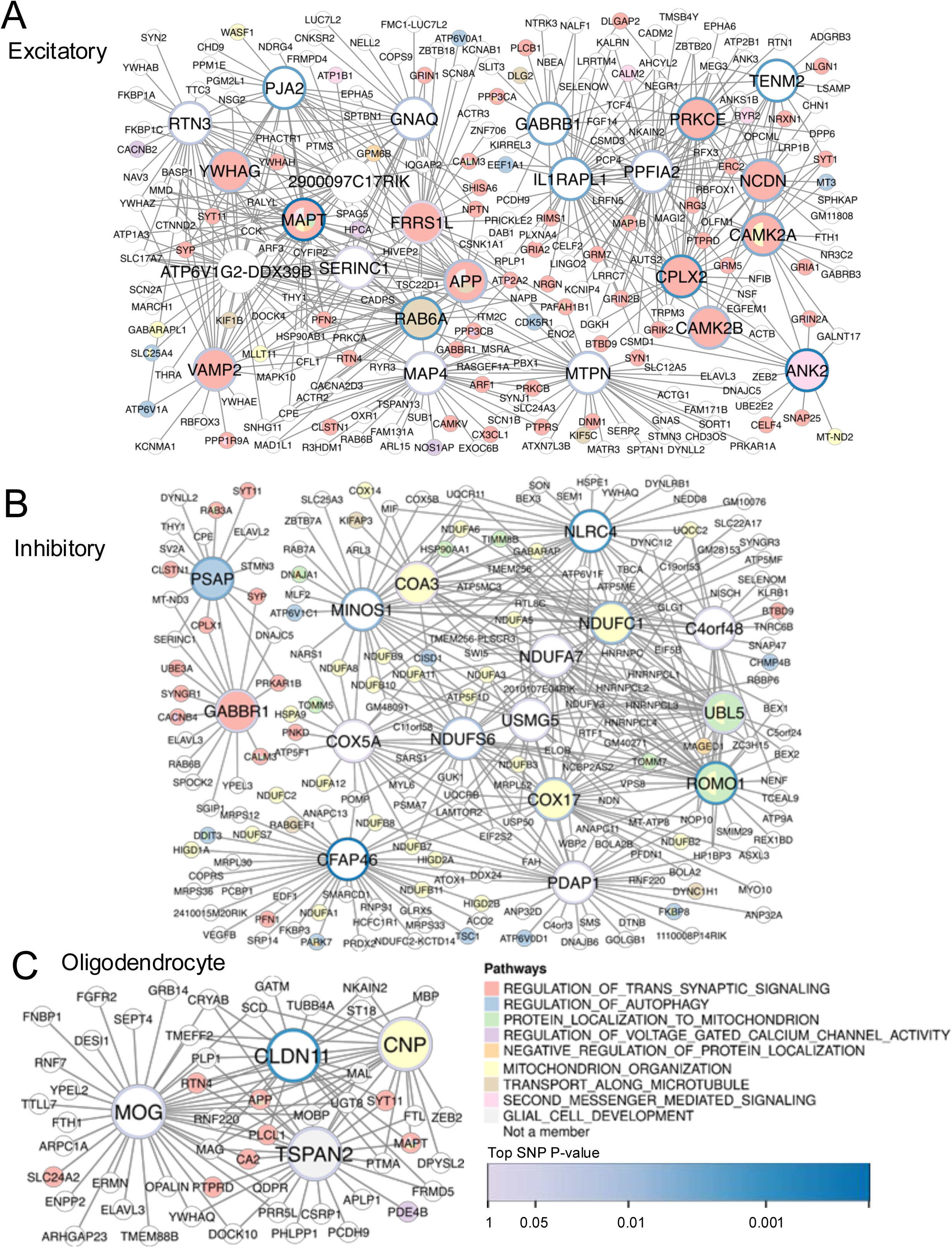
Hippocampus cell type gene regulatory networks of learning and memory pathways that interact with mTBI. Larger nodes (genes) represent key drivers of pathways and smaller nodes are the key driver neighbors. Node colors correspond to pathways, Node borders correspond to GWAS p-value of the top mapped SNP in the SNP-by-mTBI interaction term. A: Excitatory Neuron key drivers contain key drivers for synaptic signaling pathway. B: Inhibitory Neuron key drivers involved mainly in mitochondrial organization. C: Oligodendrocyte key drivers are involved in myelination and glial cell development.

Among the cortical regions, the anterior cingulate cortex contains the most KDs. For the excitatory neurons, *GRM7* (FDR=6.72e-3) and *NSF* (FDR=6.72e-3) are the top KDs for the trans-synaptic signaling pathway, whereas in inhibitory neuron *SV2A* (FDR=3.23e-5) is the top KD for the same pathway. For oligodendrocytes, *CHL1* (FDR=9.76e-3) and *TSPAN2* (FDR=9.76e-3) are the KDs for trans-synaptic signaling and glial cell development (Figure 4A). Within the prelimbic, infralimbic, and orbital cortex networks, a majority of KDs are significant for the trans-synaptic signaling pathway (Figure 4B). These include the excitatory neuron KDs *CX3CL1* (FDR=2.58e-3) and *GRIN2A* (FDR=8.37e-3), and the inhibitory neuron KDs *GRIA4* (FDR=5.29e-04) and *ATP5PD* (FDR=5.94e-03). *MOG* (FDR=0.048) is the only oligodendrocyte KD for the glial cell development pathway in the prelimbic, infralimbic, and orbital cortex networks. The frontal pole and secondary motor cortex GRNs also yielded KDs for the trans-synaptic signaling pathway, including excitatory neuron KDs *ITPR1* (FDR=9.57e-03) and *NSF* (FDR=9.93e-3). *MOG* (FDR=0.016) remains the oligodendrocyte KD for glial cell development in the frontal pole and secondary motor cortex GRNs (Figure 4C).

**Fig 4:**
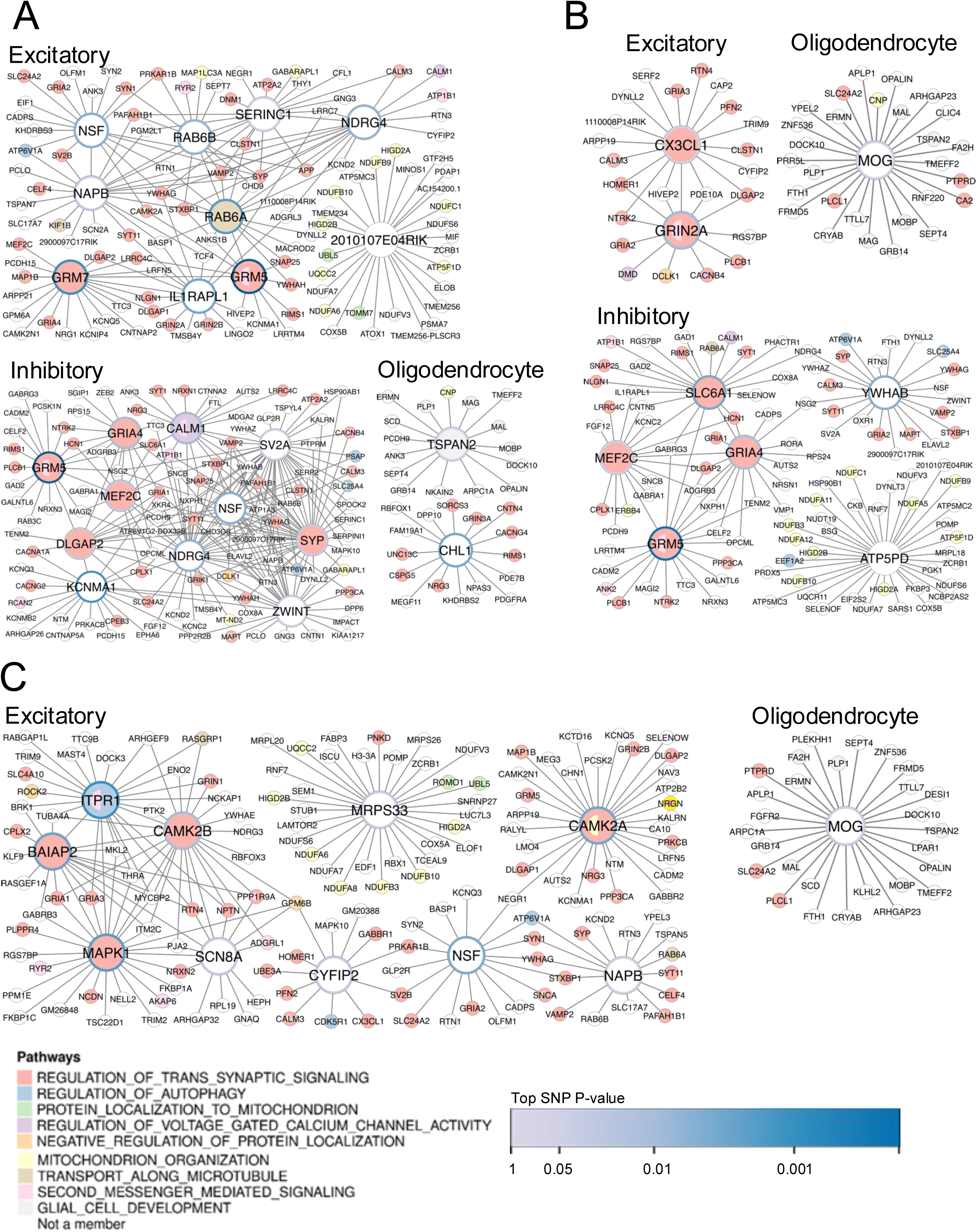
Cortex Cell Type Key Driver GRNs of learning and memory pathways. Larger nodes (genes) represent key drivers of pathways and smaller nodes are the key driver neighbors. Node colors correspond to pathways, Node borders correspond to GWAS p-value of the top mapped SNP. A: Anterior Cingulate Cortex networks. Excitatory and Inhibitory neurons contain key drivers for the synaptic signaling pathway while the oligodendrocyte key drivers additionally regulate glial cell development. B: Prelimbic, Infralimbic, Orbital Cortex networks. Both neuronal cell types drive synaptic signaling, and inhibitory neurons contain a key driver for mitochondrial organization. Oligodendrocytes contain MOG as the single key driver for glial cell development. C: Frontal Pole, Secondary Motor Cortex networks. Only excitatory neurons contain key drivers for both synaptic signaling and mitochondrial organization, and MOG remains a key driver for glial cell development.

Across brain regions, *MOG* was the most significant KD for the glial cell development pathway for the hippocampus, prelimbic-infralimbic-orbital cortex, and frontal pole-secondary motor cortex, and was the second most significant KD for the pathway in the anterior cingulate cortex. *MOG* produces a membrane protein on oligodendrocytes, and using a human leukocyte antigen linked to a MOG mouse peptide was found to improve neural deficits in a mouse model of TBI [64].

Our brain-region and cell-type specific network analysis revealed potential cell types and gene regulators of the significant biological pathways enriched for SNP-by-mTBI interaction GWAS signals, highlighting the role of excitatory and inhibitory neurons and oligodendrocytes in mTBI and their respective gene regulators of metabolic, synaptic, and glial pathways critical for neural repair and function.

### A Polygenic Risk Score (PRS) of top pathway SNPs demonstrates clinical relevance but limited utility

To evaluate the clinical relevance and potential utility of the interaction SNPs and pathways from our genetic analysis, we created a PRS for each patient based on the weighted sum of effect sizes and risk allele counts of the SNPs from the top biological pathways (Supplementary Table 8). We generated a main effect PRS based on the main-effect GWAS, and an interaction PRS based on the main and interaction effects from the interaction GWAS for learning and memory NPC3. We constructed three linear regression models using the same set of ABCD patients at baseline to test the significance of the main-effect or interaction PRS scores in predicting individual learning and memory outcomes. The null model contained injury status, age, sex, and ethnicity, while the main-effect model and the interaction model additionally considered their respective PRS scores along with the covariates (Figure 5).

**Fig 5:**
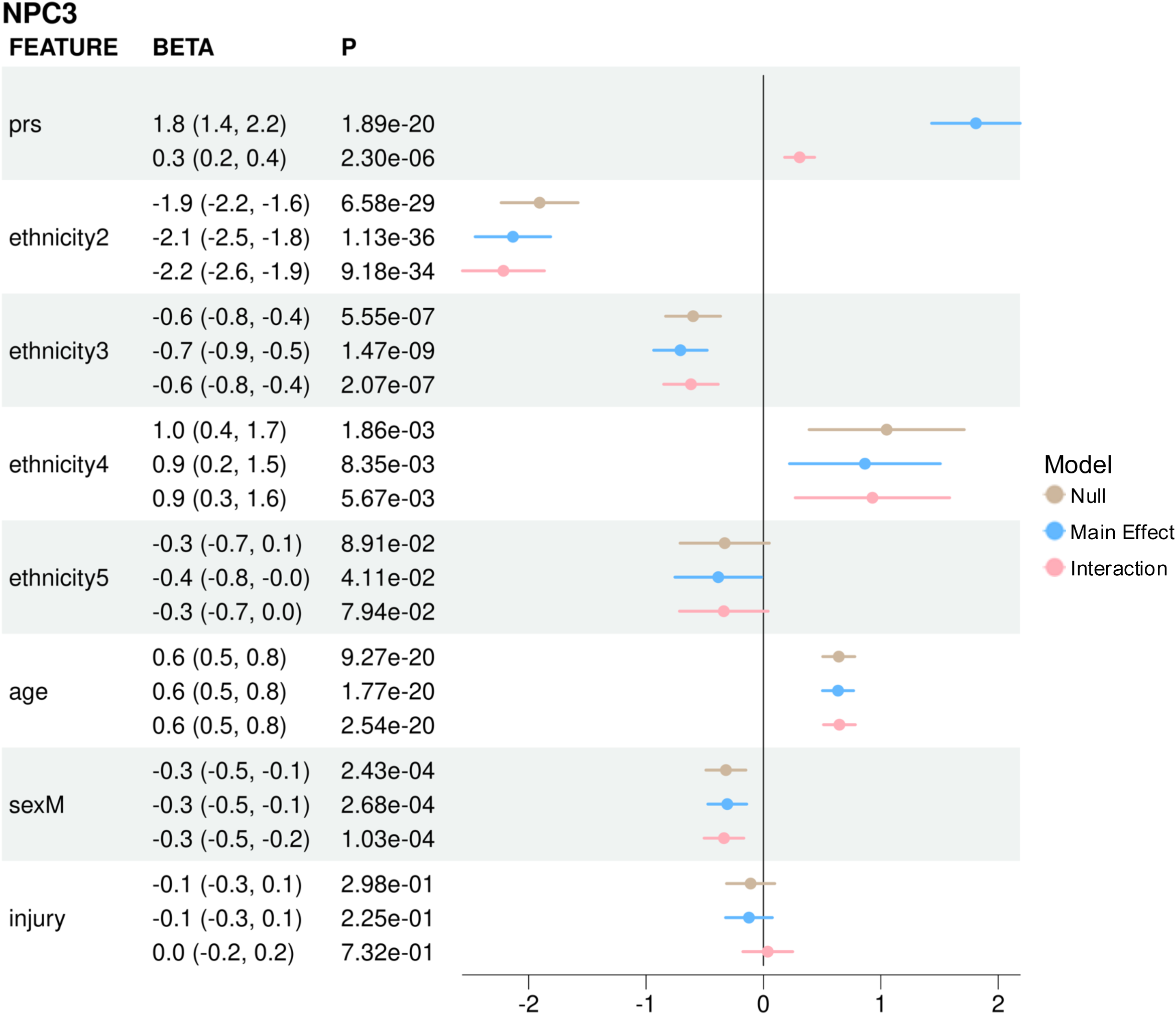
Forest plot for polygenic risk score versus other variables in linear models for NPC3. Colors are signed to the three clinical model. The x axis of the plot delineates the effect size of the linear models, accompanied by a 95% confidence interval, enabling direct comparison of the clinical models for each feature. For ethnicity, White is the reference, and 2-5 correspond to Black, Hispanic, Asian, and Other, respectively.

Across the learning and memory NIH tests (NPC3, Toolbox Picture Sequence Memory Task, Toolbox List Sorting Working Memory Task, and Rey Auditory Verbal Learning Task), the main-effect model contained the highest predictive accuracy, followed by the interaction and the null model. Model accuracy R^2^ values of the null, main-effect, and interaction models for the composite NPC3 score are 0.154, 0.204, and 0.167, respectively; for the Toolbox List Sorting Working Memory Task are 0.079, 0.106, and 0.087; for the Toolbox Picture Sequence Memory Task are 0.061, 0.082, and 0.068; for the Rey Auditory and Verbal Learning Tests are 0.070, 0.107, and 0.078 (Supplemental Figure 1). Compared to the null model, both the main-effect and interaction models significantly improved the prediction accuracy. However, the improvement for the model containing the interaction PRS over the null model is minimal and likely not clinically useful.

Effect sizes trend in the same direction across all models. The PRS term in the main-effect model indicates a 1.8-unit improvement in the NPC3 score for every unit increase in PRS, which is 6 times as large as the PRS term (0.3 units) in the interaction model. This potentially explains the higher overall accuracy in the ADD model. The average NPC3 scores across self-reported ethnic groups vary as much as 2.9, 3, and 3.1 units in the null, main-effect, and interaction models, respectively. Age and sex were also significant in these models, with a 0.6 NPC3 unit increase per year of age and 0.3 NPC3 unit decrease in males. The injury variable was not significant in any model. The significance of the PRS terms in the interaction PRS models suggests that the SNPs interacting with mTBI in the neuronal repair, synaptic signaling, mitochondrial function, and glial cell activation pathways contain genetic modifiers of mTBI recovery in learning and memory.

## Discussion

mTBI remains a leading form of brain injury, yet its effects on child neurocognitive development remain unclear, particularly given the high variability in mTBI outcomes [1–7]. With the ABCD cohort, we performed the largest gene-by-mTBI interaction GWAS to date to identify genetic modifiers of learning and memory performance affected by mTBI. Our gene-by-mTBI GWAS detected SNPs with differential effects on neurocognition based on whether the patient sustained an mTBI or OI. Despite the moderate sample size, the GWAS SNPs explain important genetic mechanisms in heterogeneous cell types and cortical regions to orchestrate neuronal and glial development and mitochondrial pathways. Furthermore, condensing these pathways into a polygenic risk score provides further insight into the clinical significance and utility for predicting neurocognitive outcomes for individuals with mTBI and orthopedic injury. This study highlights the importance of studying genetic predispositions of mTBI recovery separate from general injury recovery to elucidate the underlying biological mechanisms and key gene regulators.

Compared to previous studies on mTBI investigating genetic effects on neurocognitive and behavioral domains, the current study is a departure from the practice of using prior genetic knowledge to restrict the search for SNPs and biological pathways [10,17,22]. Here we demonstrate the advantage and power of biological discovery with data-driven approaches that surveyed all SNPs and gene ontologies to explore mTBI-and genetic-mediated neurocognitive effects and the interactions between the two. We identified novel genetic mediators of many biological pathways in mTBI pathology and brain function, such as neuronal repair, synaptic signaling, mitochondrial organization, and glial cell development. For example, we identified the top SNP for the trans-synaptic signaling pathway gene *ADRA1A*, rs138914, as a potential genetic regulator of mTBI effect on learning and memory, though previously studied in fibromyalgia [47–51]. We also report the top SNP rs10788317 that maps to *GHITM* in the mitochondrial organization pathway, which is previously associated with Parkinson’s disease gene *CHCHD2* [52,53]. We propose these genes and variants for further study in their role in influencing learning and memory. Our human genetic analysis also provided human validation of genes previously associated with poor TBI prognosis in rodent models such as *ADRA1A* and *FN1*, which were involved in trans-synaptic signaling and cell adhesion and migration pathways in our findings. Occurring in 74% of the 137 significant biological pathways, *MAPT*, which encodes microtubule associated protein Tau, is central to neuronal growth and plasticity to influence learning, and serves as a biomarker for TBI [61,65,66]. Emerging studies on the function of mitochondria in memory formation coincides with the high prevalence of mitochondrial pathways in our GWAS [67–71].

In addition to employing a pathway-based analysis on GWAS to mitigate the sample size limitation, the incorporation of scRNA-seq data of cortical and hippocampal regions from the Allen Brain Atlas elucidated the integral role of these genetic mechanisms to coordinate unique cell type regulatory pathways, drive different aspects of mTBI pathology, and control the disease’s impact on learning and memory performance. In the hippocampus, the excitatory neuron gene regulatory network focuses on synaptic signaling with key drivers like *APP, MAPT*, and *PJA2* which are known Alzheimer’s disease risk genes. These results not only align with previous studies of brain injury increasing risk for neurodegenerative disease but also propose the genetic mechanisms linking the two [59–62,72]. The inhibitory neuron GRN for SNP-by-mTBI interaction pathways mainly contains KDs involved in mitochondrial function such as *NDUFS6* and *COX5*, which have not been studied in the context of TBI. Mitochondrial dysfunction has been associated with GABA-ergic-glutamatergic neuron imbalance, which may initiate neuropsychiatric disorders [67,68,73–75]. The mitochondrial complexes 1 and 4, which include *NDUFS6* and *COX5A*, respectively, are also genetically associated with Alzheimer’s disease and memory impairment [69–71]. While mitochondrial dysfunction is a known mechanism of TBI pathology, our network analysis of the SNP-mTBI interaction pathways emphasizes the potential role of mitochondrial dysregulation at complexes 1 and 4 in inhibitory GABA-ergic neurons, specifically, to affect learning and memory. Furthermore, the oligodendrocyte network identifies glial development genes as key drivers. *MOG*, the top key driver in both hippocampus and cortical regions, has been studied for neuroprotective effects in modulating pathogenic immune functions in brain injury and stroke when administered as a partial MHC class II construct [64,76–78], and our network analysis adds further evidence supporting its functional role in modifying mTBI-induced cognitive outcomes. While previous studies focus on general gene networks without tissue and cellular context, this study emphasizes the importance of cell type heterogeneity in understanding the complexity of TBI pathology. These key drivers provide novel targets to pursue experimentally to validate their role in affecting learning and memory post-mTBI.

Lastly, we tested the clinical significance and utility of polygenic risk scores of the top genetic pathways in forecasting learning performance. Including PRS based on the main effect or interaction GWAS into clinical predictive models demonstrated increased performance accuracy over the null model without the PRS. While both terms were significantly associated with learning and memory, the increase in model accuracy over the null model without the PRS is minimal, suggesting limited clinical utility of this genetic component in predicting learning and memory. Other approaches can be incorporated to optimize the PRS, such as shrinkage, to better assess its clinical utility [79–81]. While our PRS was constructed on a small population with imbalanced proportions of different ethnic groups, it is important to test its generalizability to independent pediatric cohorts when available. We also note the strong significance in ethnicity in these clinical models. A variety of sociodemographic factors can contribute to differences in learning and memory, thus requiring deeper investigation to dissect the contribution of each of these confounders [82].

We acknowledge that this moderately-sized GWAS study has limited statistical power to detect individual genome-wide significant SNPs in the population. Despite the limitation, we found significant general agreement in the genetic effects between our GWAS in the ABCD cohort and other memory-and TBI-related GWAS in the GWAS catalog, thereby mitigating the concern. Similarly, the PRS performs well on the patients used in the GWAS study, but replication in independent and large mTBI cohorts when available, is required to confirm its generalizability to other patients. Additionally, in the current study we used chromosomal location to map SNPs to genes, but using public expression quantitative risk loci (eQTL) data would improve functional interpretation of our GWAS results. Cell-type-specific eQTL data in different tissues is currently limited but will grow and would be beneficial to include in our study to better integrate the genetic and transcriptomic layers. Regardless, the use of cell-type-specific GRNs still identifies pathways that are consistent with functions in the corresponding cell types. Lastly, we used mouse scRNA-seq data from the Allen Brain Atlas because of its granular cell type and brain region classifications. Though this atlas does not have human hippocampus data, mouse models of mTBI still provide translatable insights to humans [83–85].

Overall, our data-driven systems genetic approach explores the genetic mechanisms governing cell type processes involved in mTBI pathology regarding learning and memory. The identification of cell-type specific key driver genes and pathways provides potential targets for testing their causal and functional effects in TBI animal models and warrants future investigations into their utility and clinical translation.

## Materials and Methods

### Clinical data acquisition

Patient data was retrieved from the ABCD version 5.1 release. The ABCD study is an ongoing study including 11,878 children ages 9-10 across 21 research sites in the United States. Children undergo annual assessments for neurocognitive, behavioral, psychological, and biological endpoints. All ABCD data was accessed through the National Institute of Mental Health (NIMH) data archive (NDA).

Demographics: We used the age, sex, and ethnicity variables in our analysis. Race/ethnicity was reported as a categorical variable from 1 to 5 (1: White, 2: Black, 3: Hispanic, 4: Asian, and 5: Other/Mixed)

Injury grouping: Injury groups were defined based on 2 ABCD measures: the Ohio State Traumatic Brain Injury Screening for mTBI and the Medical History Questionnaire for OI [27,28]. We considered mTBI as patients sustaining a head injury with memory loss and/or loss of consciousness for under 30 minutes. The orthopedic injury (OI) group was assigned by parent reports on whether the child had ever visited a medical office for a bone fracture.

Neurocognitive outcome: We used neurocognitive performance scores from the NIH Toolbox Cognitive Battery (https://nihtoolbox.org/), the Rey Auditory Verbal Learning Test, and the Little Man Task. A previous analysis from Thompson et. al. derived principal component loadings to condense these variables into 3 neurocognitive principal components (NPC): NPC1 general ability, NPC2 executive function, and NPC3 learning and memory. We used the NPC3 for our analysis.

Genetic Data: We used the imputed genetic data and genetic principal components from ABCD. Data collection, quality control, linkage disequilibrium (LD), genetic relatedness filtering, and PC calculation on patient genotype data are documented in Fan et al [86]. ABCD collected ∼500,000 genetic variants for 11,666 patients using the Affymetrix Smokescreen array and imputed remaining SNPs using the TOPMED imputation panel under the GRCh38 genome build. The consortium also calculated genetic principal components using PC-AIR from the GENESIS R package to account for the multi-ancestry cohort. SNPs were pruned using GENESIS’ snpgdsLDPruning function with a 10e6 base pair sliding window and LD correlation threshold of 0.1, leaving 158,103 SNPs for PC calculation. The snpgdsIBDKING function was used to estimate kinship and identify 8,177 unrelated patients to be used in PC-AIR. The remaining unrelated individuals were projected onto the PC space using the PC loadings.

### Genome wide association study

GWAS was performed using PLINK version 1.9 [87]. We used a minor allele frequency of 0.05 to filter out rare SNPs below this threshold. Covariates included in the model were patient age, sex, injury type, and the top 20 genetic principal components. We used the kinship matrix to subset the cohort to unrelated individuals with a genetic relatedness below 0.25. We ran GWAS on all unrelated mTBI and OI patients at baseline under 2 model formulations. The first model measures the main SNP effect, and the second model includes the main and injury-by-SNP interaction effect.

### Validation of main effect GWAS in ABCD using independent working memory GWAS with large sample sizes

Though the ABCD baseline cohort of OI and TBI only contains ∼1500 patients, we validate its use for GWAS functional enrichment studies by correlating SNP effect size estimates with findings from preexisting GWAS studies on similar phenotypes. The NIH toolbox measures for learning and memory relate to working memory. We identified five GWAS studies [30–34] on verbal working memory, short term memory, and spatial working memory and correlated their summary statistics with our ABCD GWAS. We performed Pearson correlation on the effect sizes of the overlapping SNPs between ABCD and each of the five large GWAS studies.

### SNP set enrichment analysis

We used the Mergeomics method to obtain SNP set enrichment statistics using full GWAS summary statistics [45]. LD clumping was performed using Mergeomics’ marker dependency filtering, using a linkage disequilibrium r2 threshold of 0.5 to filter out redundant SNPs in high LD. SNPs were mapped to genes in pathways from Gene Ontology Biological Process from MSigDB and the GWAS Catalog [44,88] using the hg38 reference genome based on a 50 kilobase distance threshold. We calculated the enrichment of SNPs passing multiple GWAS thresholds for each biological pathway using Mergeomics’ Marker Set Enrichment Analysis (MSEA), which uses a chi-like statistic and performs a permutation test on the SNP-pathway mappings to generate true and null enrichment statistics. We used Benjamini-Hochberg correction to control the false discovery rate at 0.05 to identify significant biological pathways for the SNPs.

To further examine the association patterns of the SNPs identified in the interaction GWAS in each of the enriched pathways, we the top 20 SNPs were selected as determined by GWAS p-value for the SNP-by-mTBI interaction term. This process resulted in a collection of 345 unique SNPs, which were subsequently used to construct linear regression models on. The linear models were designed to predict cognitive test scores given the number of risk alleles a patient had for a given SNP (0, 1, 2) using *lm*(*phenotype ∼ genotype*). For each SNP, linear models were constructed for TBI and OI patients separately. The resulting p-values and effect sizes from the linear models were then used to select 41 SNPs within the enriched pathways that showed association with cognitive scores in the mTBI group but not in the OI group (Supplementary Table 8). The genotype effects in both groups are plotted to confirm and visualize the select SNPs.

### Construction of cell-type specific GRNs using scRNAseq data from the Allen Brain Atlas

We collected single cell RNA sequencing data from Allen Brain Atlas and classified brain regions and cell types based on the Allen Brain Institute’s published cell taxonomies [26]. We used Single Cell INtegrative Gene regulatory network inference (SCING) to construct global GRNs for each cell type [58]. From scRNA-seq data, SCING first mitigates cell sparsity by pseudo-bulking similar cells using Leiden clustering on the K-nearest neighbor graph for each cell type. To infer regulatory relationships SCING trains gradient boosting machines to predict target gene expression based on their co-expressed genes and use feature importance measures of the feature genes to weigh directed network edges. For robustness, SCING bootstraps 100 subsets from the pseudo-bulk data, performs the GRN inference step for each subset to yield 100 intermediate GRNs, and aggregates recurring network edges among the intermediate GRNs to produce a final consensus GRN. We performed SCING for each of the cortical and hippocampal cell types from the 10X chromium scRNA-seq data in the Allen Brain Atlas.

### Key Driver Analysis (KDA) of biological pathways

Genetic mechanisms underlying TBI consequences of learning and memory were identified using KDA on each of the cortical and hippocampal cell type GRNs [45]. KDA uses the GRN topology to identify hub genes with high connectivity to gene sets of interest, in this case enriched biological pathways from MSEA analysis of the SNP-by-mTBI interaction GWAS. It identifies hub gene neighborhoods enriched for the gene sets through a chi-like statistic, and measures significance of the enrichment based on the null distribution of enrichment scores generated from permuted networks. The significant hub genes at FDR<5% are proposed as the key drivers of the mTBI-interacting pathways. We performed KDA on normal mouse hippocampus and prefrontal cortex cell type GRNs.

### Polygenic risk score modeling

We calculated the polygenic risk score using the most significant SNPs from each MSEA pathway (Supplementary Table 8). We used PLINK version 1.9 to calculate the PRS [87]. We calculated PRS based on 2 GWAS models: one including the main and interaction terms (interaction PRS) and one with only the main SNP effect term (main-effect PRS). We calculated PRS scores for the same patients used in the GWAS. For predictive models, we built linear regression models on the NPC3 learning and memory variable, as well as the Toolbox Picture Sequence Memory Task, Toolbox List Sorting Working Memory Task, and Rey Auditory Verbal Learning Task. We included covariates of age, sex, and ethnicity in the model and compared the model performance when including the main-effect PRS, interaction PRS, or no PRS (null model). The models were structured as follows:

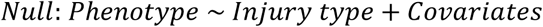

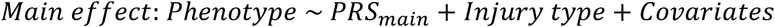

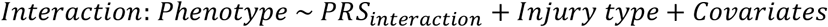

PRS represents the polygenic risk score calculated under the main-effect model or the interaction model. Injury type indicates the presence of mTBI, coded as 1, as opposed to OI, coded as 0. The covariates include self-reported race/ethnicity (represented as a factor from 1-5 for White, Black, Hispanic, Asian, and Other/Mixed), age in months, and sex (M or F). The reference groups for the models were 0 (OI) for injury, 1 (White) for ethnicity, and F (female) for sex.

### Visualization

All plots were generated in R version 4.1.0, and all networks were generated with Cytoscape [89]. We visualized GRNs the direct neighbors of the significant key drivers in each cell type.

### Data and code availability

All data pertaining to ABCD is available through National Institute of Mental Health Data Archive. Data analysis scripts can be found in our Github repository https://github.com/XiaYangLabOrg/ABCD_GWAS or by contacting the lead author.

### Author Contributions

Michael Cheng, Xia Yang, Jeffrey Max, and Florin Vaida conceptualized the study design and methodology. Michael Cheng, Joanna Jacobus, Emily Troyer, and Wenjing Meng curated the data. Joanna Jacobus and Jeffrey Max created the positive and negative control sets from the GWAS Catalog. Michael Cheng and Melody Mao contributed to the data analysis. Michael Cheng, Melody Mao, Wenjing Meng, Emily Troyer, and Xia Yang wrote the original draft. All authors reviewed and edited the final draft.

## Data Availability

All ABCD data was accessed through the National Institute of Mental Health 418 (NIMH) data archive (NDA)

## Supporting Information Captions

**S1 Table: NPC3 main effect GWAS summary statistics for suggestive SNPs (p<=1e-5).**

Columns include SNP id, chromosome and base pair location, effect size, and p-value.

**S2 Table: Correlations of summary statistics between the main effect NPC3 GWAS and the working memory and negative control GWAS studies.**

**S3 Table: NPC3 interaction GWAS summary statistics for suggestive SNPs (p<=1e-5).** Columns include SNP id, chromosome and base pair location, main-effect and SNPxInjury effect sizes and p-values.

**S4 Table: MSEA results for GWAS catalog positive and negative control traits.** Columns include GWAS trait, enrichment score, p-value, number of genes, number of SNPs, FDR, top overlapping SNPs, top mapped genes, top SNP interaction GWAS p-values, and positive/negative control set categorization.

**S5 Table: MSEA results for NPC3 interaction GWAS on the full GWAS catalog.** Columns include GWAS trait, enrichment score, p-value, number of genes, number of SNPs, FDR, top overlapping SNPs, top mapped genes, top SNP interaction GWAS p-values. Traits highlighted in blue are significant enrichments (FDR < 0.05).

**S6 Table: MSEA results for NPC3 interaction GWAS on the GO-BP.** Columns are the same as Supplementary Table 5, except replacing GWAS catalog trait with GO-BP pathway. Traits highlighted in blue are significant enrichments (FDR < 0.05).

**S7 Table: Cell Type KDA results.** Columns include key driver gene, brain region, cell type, key driver pathway, key driver enrichment and FDR, top mapped SNPs to key driver, interaction GWAS p-values of the top mapped SNPs.

**S8 Table: SNPs used for PRS.** Columns include SNP ID, chromosome, mapped gene, significant GO-BP pathways, main-effect and interaction GWAS effect sizes and p-values, effect size, p-value, and FDR in the unadjusted linear models on NPC3.

**S1 Figure: Forest plot for clinical PRS models on individual neurocognitive test scores.**

